# New SIR model used in the projection of COVID 19 cases in Brazil

**DOI:** 10.1101/2020.04.26.20080218

**Authors:** Augusto S. Freitas, Silvio S. Lacrose Sandes, Leonardo S. Silva

## Abstract

In this work, we proposed a variant of the SIR model, taking as based on models used to describe the epidemic outbreak in South Korea and Portugal, to study the COVID-19epidemic curve in Brazil. The model presented here describes with reasonable agreement the number of COVID-19 cases registered in Brazil between February 26 and May 18, 2020 based on the hypothesis that there a large number no notified cases (11 to 1) and variation in contagion rate according to social isolation measures and greater or lesser exposure to the virus (highest rate in beginning from epidemic). To this end, we introduced an exposure factor, called β_1_/β_2_, which allows us to describe the influence of factors such as social isolation on dispersal from disease. The results also corroborate a phenomenon observed in countries that registered a high growth in cases in short period of time, to example of Italy, Spain and USA: if isolation measures are imposed late, the total number of cases explodes when the epidemic is approaching from peak, which implies a higher exposure rate in the first days of case registration. From the data collected, we made the linear adjustment of the infected curve according to the time between the 41st and 74th days since the official notification of the first case and we obtained a high infection rate in the period, close to 4.0. The result indicates that the relaxation of social exclusion measures contributed to the high increase in cases in the period. This result reinforces the adoption of the model that differentiates unexposed susceptible from those that are most exposed to contagion. The model also predicts that the peak epidemic outbreak in Brazil, based on the number of cases, will occur around in late May and early June.

## 1. INTRODUCTION

Since the outbreak of the disease COVID-19 in the city of Wu Han, China, in just over two months the epidemic spread rapidly around the world, until that on March 11, 2020, the World Health Organization (WHO) declared the COVID-19 pandemic (ZHANG *et al*., 2020). Until May 18, 2020 the number of cases in the world had already exceeded 4.6 million, with approximately 300.000 confirmed confirmed deaths.

With the explosion in number of cases from COVID-19 in Brazil and worldwide, to understand how it happens the evolution of the number of cases is important to evaluate the measures taken until the moment were effective, as well to determine the next actions to be executed in order to prevent the collapse of the country’s health system. In 1927, W. O. Kermack and A. G. McKendrick (1927) created a model that considers a fixed population with only three compartments: sensitive, S(t); infected, I(t), and removed, R(t) - SIR (Susceptible-Infected-Recovered) - for the first time, in order to study the evolution of epidemic processes and how the spread of infectious diseases over time. Other models were created along the history, for example one developed by Daniel Bernoulli in 1790 to study the variola epidemic (HERBEN, 2000).

The SIR model allows to project the evolution of an infectious disease in a population in agreement with their state transition rates and their initial conditions S(0), I(0) and R(0) and it gets, with reasonable precision, to describe quantitatively, for instance, the time interval from the first contagion until the number of infected people reaches the epidemic peak, important information that allows governments to develop actions to reduce the number of having infected or, at least, extend the time in that peak of infections will be reached (SCHIMIT, 2010; JO, 2020). However, there are variants of the SIR model that can be applied in situations in which another variables are important for description how the infection spreads in a population of S susceptible individuals, to example as was done in Teles’ work (2020) adapting the SIR model to describe the behavior of the coronavirus infected curve in Portugal.

In this work, we propose a variation of the SIR model, taking initially described by JO *et al*. work (2020), to describe the projection of cases of COVID-19 in a qualitative and quantitative way in Brazil since the first day in wich cases were confirmed, February 26, 2020, until May18, 2020. For this, we will use what here we will call SIER model, Susceptible-Infect-Exposed-Recovered, that is, a model that also considers that there exposed individuals, represented by the *E*(*t*) function, which have a higher probability of infection than average populational. In the following sections, we will present the model and the method used to solve the differential equations originating from it; then we will present the results obtained, following by the final considerations.

## 2. MODEL AND METHODS

In this work a variant of the S. I. R. model - Susceptible, Infected and Removed (recovered and/or died), with fixed number for the population (normalized, *N*=1), without taking into account the amount of births/deaths, but taken into account that, in the universe from susceptible individuals, S(t), there is a smaller number of individuals E(t), that are those susceptible, however more exposed to infection, to example of people that be not able to adhere the complete social isolation or professionals of health, that are in frontline from caring of those infected. The model takes into account that a S(t) - E(t) number of individuals can be infected at rate β_1_ and a E(t) number of individual can be infected a rate β_2_>β_1_, once the exposed individuals have larger probability to contract the virus and infect other individuals. The functions that represent the susceptible individuals, S(t), infected, I(t), exposed, E(t), and removed/recovered, R(t), are link with their respective rates of temporal variation from the system of differential equations (to a time t > 0):

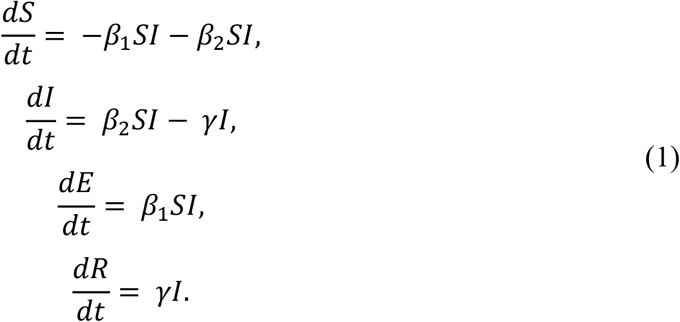

where γ is the average recovery rate (1/γ is the period during which the infection continues), and the functions S(t), I(t), E(t) and R(t) obeying the following conditions:

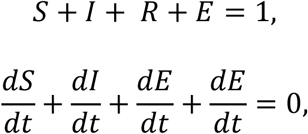

where

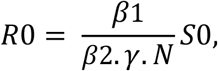

is the effective contagion rate (reproduction number) (with *N* = 1).

In the S.I.R. conventional, at the beginning of the epidemic, the number of susceptible individuals approaches the unit (normalized population), S ≈ 1, therefore the second of equations (2), becomes:

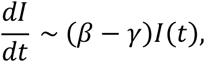

whose approximate solution is

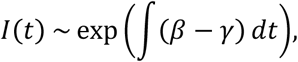

being the number of effective reproduction, in a time interval *T*, given by 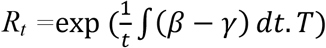 (JO et al, 2020). It is important to note that this means that the reproduction factor changes over time, growing exponentially to low t, reaching a peak (at the peak of infection) and decreasing more slowly after that, as predicted by the work of JO et al. (2020). The faster the reproduction factor falls, the faster the epidemic will be controlled, an important data for the adoption of measures that will mitigate the effects of the pandemic in the state and that needs to be taken into account by health authorities.

This model is appropriate for diseases that spread quickly and give immunity to survivors, to example as the conventional SIR (SCHIMIT, 2010), but can, without loss of generality, be used to describe the installation of agricultural pests or spread of computer virus. In most varied lattices topologies, for instance, since the propagation processes, of the disease or information, are very similar (HASTINGS, 2003; PACHI, 2006). There are other models that take into account differences between the propagation mechanisms or even larger complexity in the individuals’ interaction, but they will not be treated in this work.

The Eqs. (1) are of difficult analytical resolution, what takes to approaches or it looks for numeric treatment to arrive to the curves that represent S(t), I(t), E(t) and R(t). To this, a computational routine was developed for language Octave with the intention of obtain the numeric solutions for that model and to apply it to study spread of COVID-19 in Brazil, without considering the differences of each place. Octave is a software of free distribution, following the patterns of distribution of the General Public License (GNU), with several functions for resolution systems of differential equations for instance (SCHERER, 2005). Here, we suppose that all individuals belonging to population have equal contracting/transmitting probability the disease in certain time interval (KEELING, ROHANI, 2008; NEWMANN, 2002).

## 3. RESULTS AND DISCUSSION

Predictions from the model proposed in this work are useful only for describing a time interval in which the Brazilian health system is more likely to collapse due to the epidemic peak. We must take into account that the differential equations used are not linear, therefore highly susceptible to small changes in level or occurrence, and that no mathematical model incorporates all the variables that can influence the behavior of the epidemic curve, bringing a simplified picture of spread of the epidemic based on simple interactions between the susceptible, and these models do not incorporate a possible temporal variation of the reproduction factor.

The presented results will be analyzed according with the following premises: **i)** The number of cases is, at least, eleven times greater than that officially registered (WALKER et al., 2020); **ii)** The rate of contagion doesn’t vary with time and is the same for all individuals; **iii)** The relationship among β_1_ and β_2_ parameters, specifically the ratio β_1_/β_2_, is the important coefficient for analysis from impacts of high exposure to virus and its consequences in which concerns the spread of infection by population; iv) The absence of politics of social isolation (high exposure from relatively large portion of population) implies more, in our analysis, a greater number of cases than a significant change at time when the epidemic curve reaches the peak maximum.

The graph in Fig. 1 shows the behavior of the curves of infected and exposed individuals (that still didn’t contract the disease) in function time, in agreement with the model proposed in this work. The curves show a typical epidemic pattern, in a qualitative way, however there is slight dependence between the date which the peak of infection occurs and total percentage of infected (number of accumulated cases) at the peak of infection: the larger contagion factor β_1_/β_2_, the larger the total number from infected and if the total number of infected is greater (especially in the epidemic peak), the impact on health system is considerably greater (taking into account the same rate of contagion in two cases).

**Figure 1.**
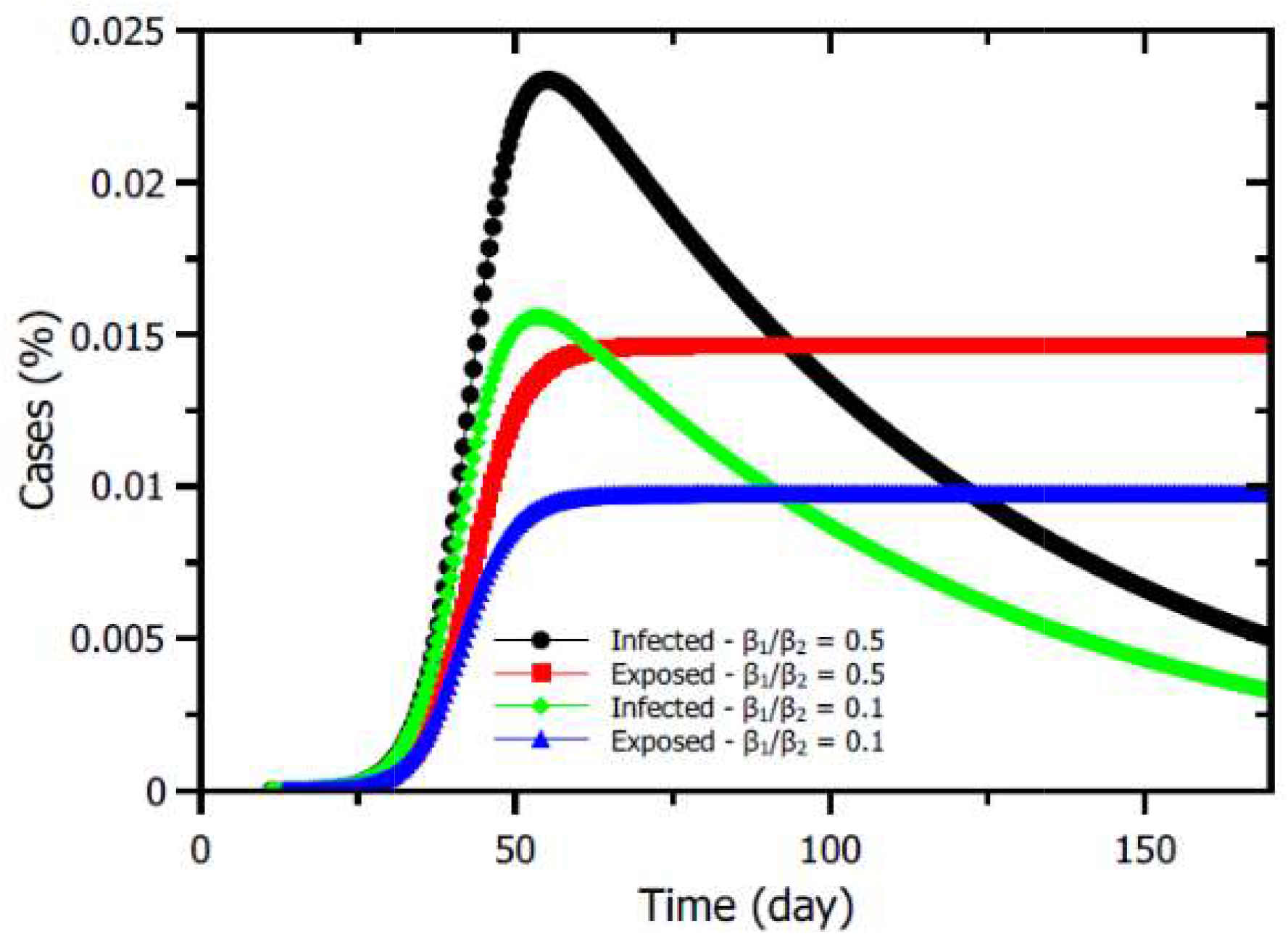
Curve showing the number (percentage) of infected and exposed individuals in time function to different values from exposure factor, β_1_/β_2_.

It’s observed that there is no significant difference in the number of cases at beginning from epidemic, independent of greater number or not from highly exposed susceptible individuals. This characteristic has been observed in several countries, especially in those where politics of social isolation were not initially imposed. In the beginning of the outbreak, governments that did not adopt isolation politics because, apparently, the epidemic outbreak had lost force, were eventually forced to adopt them, and once the cases number exploded after a short period and this is exactly the behavior visualized in the epidemic curves shown in Fig. 1.

The graphs from figures 2 and 3 shows the epidemic curve for different scenarios, with slightly different β_1_/β_2_ ratios, as well as different contagions. In Figure 1 we have the variation in number of infected individuals (percentage of infected in relation to the whole Brazil population (210.147.125 people - IBGE, 2019) along little more than sixty days as time function.

**Figure 2.**
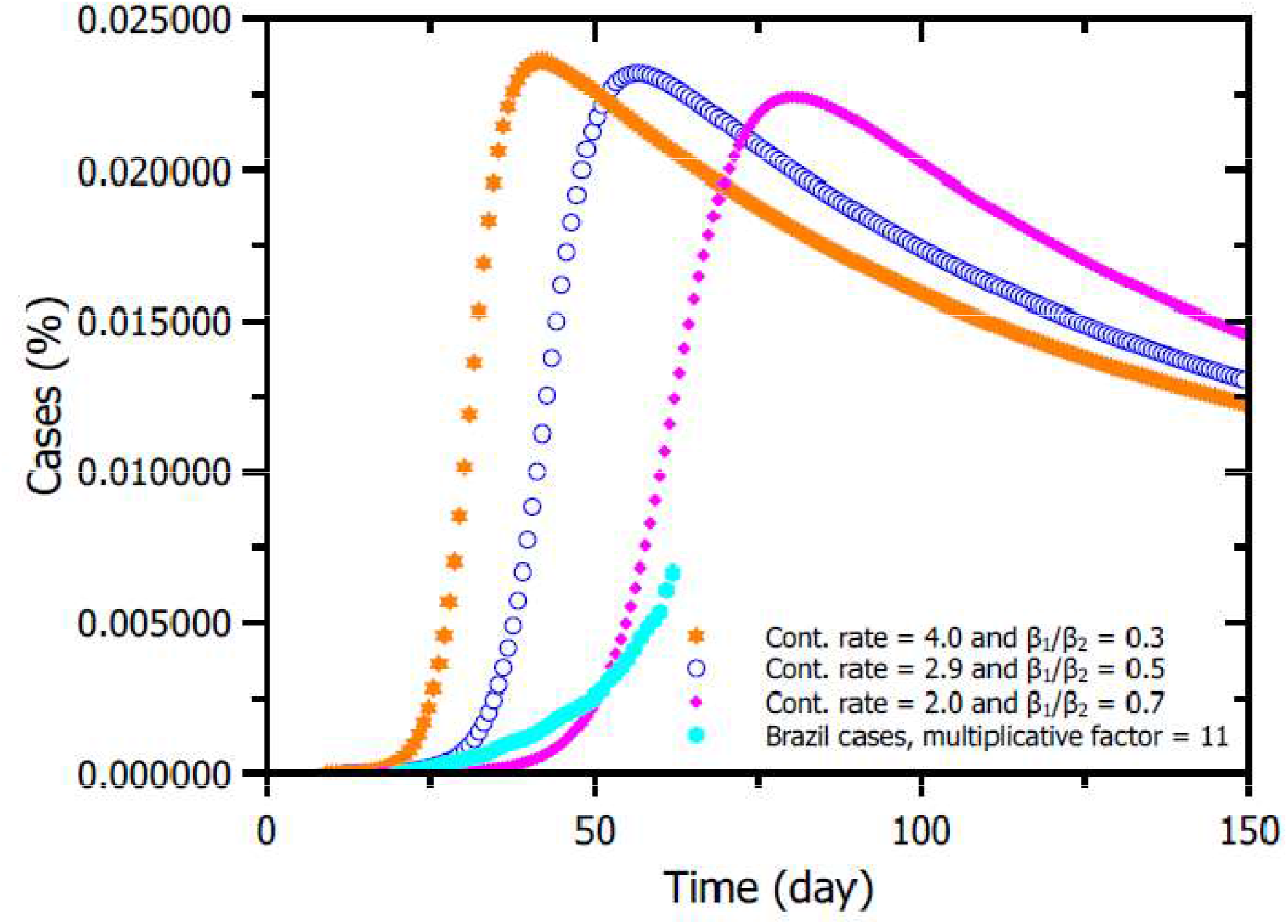
Epidemic curves for several scenarios, with different contagion rates and exposure factors. Using the hypothesis that there are eleven cases for each case officially notified in Brazil (WALKER et al., 2020). The curve that best fits the case number registered in the country between February 26 and May18, 2020 is to contagion factor equal to 2.0. The graph shows, taking into account the contagion rate curve 2.0, that the epidemic peak will be reached around (late May and early June). Data available at: Coronavirus Brazil, https://covid.saude.gov.br/.

**Figure 3.**
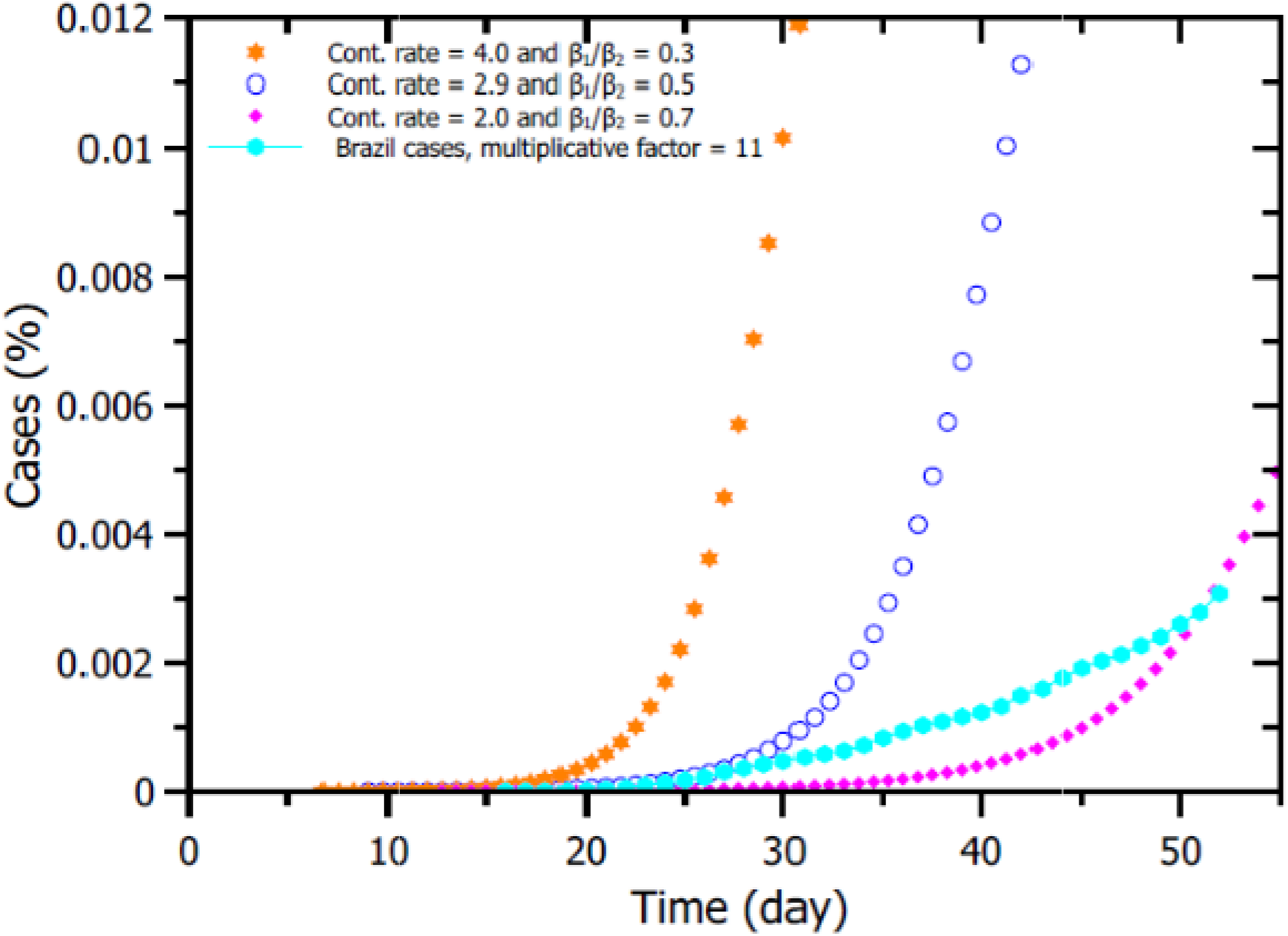
Curves with the percentage of infected (in relation to the total Brazilian population) in an interval of 55 days.

The epidemic curve in Fig. 2 present a compatible pattern with infection models with exponential growth during the initial period, change in the concavity of curve that describes the number of infected, I(t), until reaching the peak, for a given percentage of cases, and the decrease from accumulated amount from registered cases. Interesting to observe that, using the hypothesis that there are eleven cases for each officially notified case in Brazil (WALKER *et al*., 2020), the curve that best fits the number of cases registered in the country between February 26 and May 18, 2020 is that to contagion factor is equal to 2.0. It’s also observed that one that more influences in the fast initial virus spread is the contagion rate, serving the exposure factor β_1_/β_2_ as a way to differentiate the speed with which the epidemic will spread taking into account a same contagion rate. For instance, if the contagion rate is high, even with a low β_1_/β_2_ exposure factor, the epidemic spreads quickly. The graph in Fig. 2 display, taking into account the contagion rate curve 2.0, that the epidemic peak will be reached around late

May and early June, but is important have caution about this data, once Brazil is a country from continental dimensions, with important regional differences that may indicate that each state will reach the peak in different dates.

In Fig. 3 have the percentage data of infected people over thirty days, considering the first thirty days starting from the first case registered in Brazil (February 26, 2020, day 1) until May 18, 2020. It can be observed in this graph that the number of cases (hypothesis of eleven for each registered case) in Brazil increase with a contagion rate close in 2.0 in the first thirty days from epidemic outbreak, dropping to 2.0 after thirty days, indicating that measures of social isolation adopted in the country, even if partially, served to reduce the speed of spread from virus, even with different rates of beta exposure, which indicates that the ideal is to combine the rates of contagion and exposure this way we have a real picture how the epidemic is spreading.

It’s evident that contagion isn’t fixed in time and depends strongly on isolation measures: In the beginning from pandemic, with many exposed individuals, the contagion is high and can remain so (or even increase) if no measure of social isolation is adopted, falling continuously with time, because a combination of isolation measures with factors such as climate and seasonal diseases. For instance, in winter, flu-like syndromes are more common, which ends up taking more people to hospitals (increased exposure rate). Going to hospitals, these people are subject to contact with other individuals infected by coronavirus, fact that increases the contagion chance both for those have another type of comorbidity and for their companions.

The real data from pandemic is very underreported, a rising carried out by Open Knowledge Brasil (2020), organization that act in the area of transparency and openness from public data, in evaluation that considered the content, format and level of detail from information published on the portals from state governments and federal government; 11 Brazilian states do not publish minimum data, in which 90% of the evaluated states, until April 3, 2020, still did not publish enough data to accompany the spread from Covid-19 pandemic across country, including the federal government. Like this, any scenario drawn by mathematical systems based on government data can’t represent the real situation of the pandemic in Brazil.

In addition, until April 2, 2020, more than 25 thousand tests for COVID-19 were still waiting for results, a number three times higher than the number of confirmed cases until that date. Considering that the Ministry of Health from Brazil recommends testing only severe cases, considering that tests are still lacking even for severe cases, added to fact that large part from population is asymptomatic for COVID-19, we noticed that the number of underreported cases in Brazil it is extremely underestimated, which reinforces the hypothesis adopted here, based on the study by Walker *et al*. (2020) eleven cases for each made registration.

The analysis of the behavior from epidemic curves shown in the graph in Fig. 3 ensures the validity of one of our hypotheses, which the contagion rate in Brazil is between 2.0 and 3.0. If no measure of social distancing or decrease in exposure the contaminated was adopted, the contagion rate could be higher than 3.0.

The curve adjustment shown in the graph in Fig. 4 indicates a high rate of contagion, close to 4.0, and explains the explosion of cases registered in the country from the second/third week of April to the third week of May. This high increase in the number of cases as recorded can be associated with the relaxation of social withdrawal measures after the Easter holiday, especially in states whose isolation rate was below the 70% recommended by WHO. According to data published by the newspaper Estadão (2020), the social isolation index in Brazil reached its peak on March 22, 2020, having since fallen to something close to 40% on May 16, 2020. The drop in the isolation rate is the most likely explanation for the high rate of contagion in this period.

**Figure 4.**
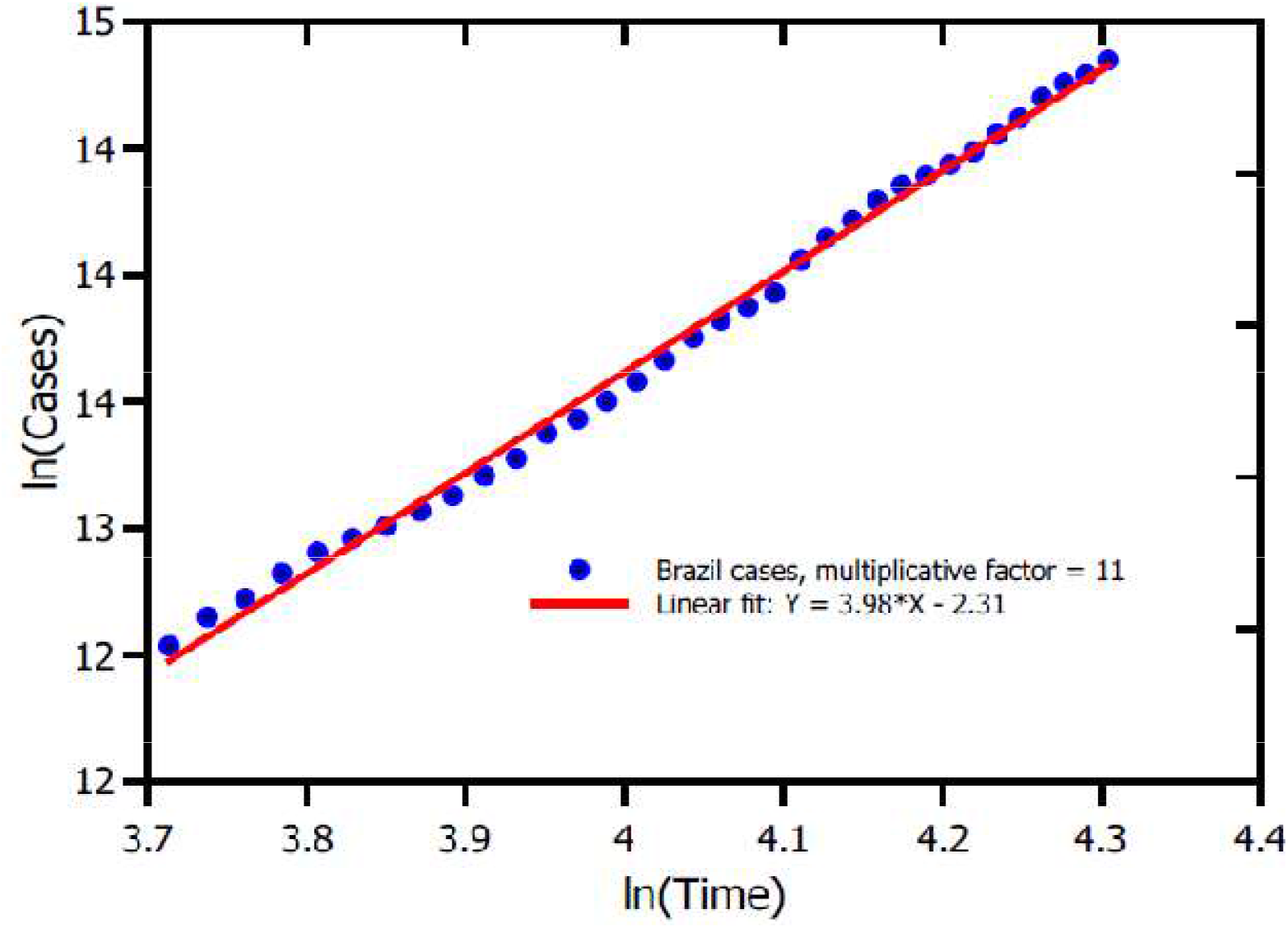
Graph with curve fit for COVID-19 cases registered in Brazil (considering multiplicative factor = 11) between the 41st day after the first record and the 74th day. The adjustment indicates an increase in the speed of spread of the disease in the country, with the next step factor being 4.0, which is the value of the variation in the recovery. One possible explanation for these high rates of contagion is the “loosening” of social withdrawal measures after the Easter holiday.

Is important to express that the curves modeled by the S.I.R. model not consider crucial factors that can lead to even worse panoramas, such as, for instance, the virus is already present in slums in Brazil, where more than 13 million people live, according to the Locomotiva Institute (2020). In these places, families live in just one room, being impossible to separate risk groups, such as senior, from people who need to go out to work and return home, increasing the risk of contamination. In many of these communities, there are no basic sanitation conditions, impending simple processes such as hand hygiene frequently, as recommended by authorities.

## 4. FINAL CONSIDERATIONS

In summary, the model presented here describes with reasonable agreement the number of cases from COVID-19 registered in Brazil between February 26 and May 18, 2020 with based on hypothesis that there area large number of underreported cases (11 to 1) and variation in the contagion. The rate of contagion according to measures of social isolation and greater or lesser exposure to the virus (highest rate at the beginning of the epidemic). The results also corroborate with a phenomenon observed in countries that registered a high growth in cases in a short period of time, such as Italy, Spain and the USA: if isolation measures are imposed late, the total number of cases explodes in way epidemic is approaching from maximum peak, which implies a higher exposure rate in the first days of case registration. From the collected data, we made the linear adjustment of the infected curve according to the time between the 41st and 74th days since the official notification of the first case and we obtained a high infection rate in the period, close to 4.0. The result indicates that the relaxation of social exclusion measures contributed to the high increase in cases in the period. The model also predicts that the peak epidemic outbreak in Brazil, based on the number of cases, will occur around in late May and early June.

## Data Availability

The data that support the findings of this study are openly available in Painel Coronavirus at https://covid.saude.gov.br/

